# Changes in Characteristics of Drug Overdose Death Trends during the COVID-19 Pandemic

**DOI:** 10.1101/2021.02.01.21250781

**Authors:** Catherine DiGennaro, Gian-Gabriel P. Garcia, Erin J. Stringfellow, Sarah Wakeman, Mohammad S. Jalali

## Abstract

**Background:** Reports analyzing drug overdose (OD) mortality data during the COVID-19 pandemic are limited. Outcomes across states are heterogenous, necessitating assessments of associations between COVID-19 and OD deaths on a state-by-state level. This report aims to analyze trends in OD deaths in Massachusetts during COVID-19.

**Methods:** Analyzing 3,924 death records, we characterize opioid-, cocaine-, and amphetamine-involved OD mortality and substance co-presence trends from March 24-November 8 in 2020 as compared to 2018 and 2019.

**Results:** OD deaths involving amphetamines increased by 85% from 2019 to 2020 (61 vs. 113; *P*<0.001) but were steady from 2018 to 2019. Heroin’s presence continued to decrease (341 in 2018, 247 in 2019, 157 in 2020; *P*<0.001); however, fentanyl was present in more than 85% of all OD deaths across all periods. Among OD deaths, alcohol involvement consistently increased, present in 250 deaths in 2018, 299 in 2019 (*P*=0.02), and 350 in 2020 (*P*=0.04). In 2019, 78% of OD decedents were White and 7% were Black, versus 73% and 10% in 2020 (*P*=0.02).

**Conclusion:** Increased deaths involving stimulants, alcohol, and fentanyl reflect concerning trends in the era of COVID-19. Rising OD death rates among Black residents underscore that interventions focused on racial equity are necessary.

## Introduction

The COVID-19 pandemic has revealed deep cracks in the U.S. health care system, social support structures, and economy. Meanwhile, another epidemic is increasingly deadly; over 90,000 people are estimated to have died of a drug overdose (OD) in 2020, as compared to over 70,000 in 2019 (National Center for Health Statistics, 2021), the largest increase since at least 1999. While opioid-involved deaths constitute the majority of OD deaths, ODs involving cocaine and stimulants have risen (O’Donnell, Gladden, Mattson, Hunter, & Davis, 2020), and alcohol consumption has increased (Pollard, Tucker, & Green, 2020). The overdose crisis has evolved into one of the leading causes of death over the last 20 years, and the COVID-19 pandemic may have contributed to its acceleration over the last 15 months. Acknowledging the potential of COVID-19 to thwart harm reduction and treatment access, experts predicted that an OD surge would be exacerbated by isolation, economic instability, and volatility in opioid tolerances and supply (Wakeman, Green, & Rich, 2020).

Preliminary results in California (through April 18) and Indiana (suspected ODs through July 24) show increases in daily OD deaths (Glober et al., 2020; Rodda, West, & LeSaint, 2020), and nationally, overdoses appear to have increased by nearly 60% in May 2020 as compared to May 2019 (Friedman & Akre, 2021). States’ OD and COVID-19 trajectories differ greatly (Jalal et al., 2018), necessitating assessments of the relationship between the pandemic and ODs on a state-by-state level. In this report, we analyze drug OD mortality trends in Massachusetts during the COVID-19 pandemic as compared to 2018 and 2019, using the latest available mortality data from the state.

## Methods

### Samples and Measures

We obtained individual-level data for deaths recorded from 2018 to 2020 from the Massachusetts Registry of Vital Records and Statistics. We used ICD-10-CM codes to identify ODs involving opioids, cocaine, or psychostimulants as a cause of death (opioids: T40.1, T40.2, T40.4, T40.6; cocaine: T40.5; psychostimulants: T43.6) and restricted the study period to March 24 (stay-at-home order enacted) to November 8. At the time of our analysis, data after November 8, 2020 were considered provisional.

We extracted all substances present at the time of death from the text of medical examiners’ final reports, sorting substances into seven relevant categories: prescription opioids (hydrocodone, hydromorphone, oxycodone, oxymorphone, codeine, dihydrocodeine, levorphanol and tramadol), heroin, fentanyl, fentanyl analogs, alcohol, benzodiazepines, amphetamines (including methamphetamine), and cocaine. We also obtained COVID-19 case fatality rates across all Massachusetts counties from the COVID-19 Data Repository at the Center for Systems Science and Engineering (CSSE) at Johns Hopkins University, which is publicly accessible.

### Analyses

To identify changing trends in OD deaths from 2018-2019, we assessed differences in distributions of age and daily OD death counts using the Mann-Whitney U test and differences in sex, manner of death, race, and substance variables using Pearson’s Chi-Square Test of Independence. In a follow-up to our main analysis, we further assessed the relationship between COVID-19 severity and OD deaths by computing Pearson’s correlation between COVID-19 case fatality rates and the percentage change in OD deaths per 100,000 people at the county level.

Analyses were performed using R version 4.0.3. This study was deemed exempt from review by Mass General Brigham’s institutional review board. The analysis was not pre-registered, and the results should be considered exploratory.

## Results

During the March 24 to November 8 analysis period, there were 1305 recorded OD deaths in 2018, 1298 deaths in 2019, and 1321 deaths in 2020. Population characteristics and results are described in Table 1.

**Table 1:** Characteristics of overdose deaths in Massachusetts

| Characteristic | No. (%) |  |  | P-value<sup>a</sup> |  |
| --- | --- | --- | --- | --- | --- |
|  | (A) | (B) | (C) | (A) vs. (B) | (B) vs. (C) |
|  | 2018<br>Mar 24-Nov 8 | 2019<br>Mar 24-Nov 8 | 2020<br>Mar 24-Nov 8 |  |  |
| **Total overdose deaths** | 1305 | 1298 | 1321 |  |  |
| **Daily overdose deaths [95% CI]** | 5.7 [5.4-6.0] | 5.7 [5.4-6.0] | 5.8 [5.5-6.1] | 0.75 | 0.61 |
| **Sex** |  |  |  |  |  |
| Male | 953 (73) | 969 (75) | 971 (74) | 0.37 | 0.53 |
| **Age [95% CI], years** | 41.2 [40.6-41.9] | 41.6 [40.9-42.3] | 42.7 [42.0-43.3] | 0.40 | 0.02* |
| **Race/Ethnicity** |  |  |  | 0.10 | 0.02* |
| White, non-Hispanic | 1044 (80) | 1013 (78) | 970 (73) |  |  |
| Black, non-Hispanic | 66 (5) | 90 (7) | 130 (10) |  |  |
| Hispanic | 151 (12) | 167 (13) | 181 (14) |  |  |
| Native American/Alaska Native/other | 27 (2) | 17 (1) | 28 (2) |  |  |
| Asian, non-Hispanic | 15 (1) | 11 (1) | 11 (1) |  |  |
| Unknown | 2 (0.1) | 0 | 1 (0.08) |  |  |
| **Manner of death** |  |  |  | 0.70 | 0.50 |
| Accident | 1270 (97) | 1261 (97) | 1274 (96) |  |  |
| Suicide | 22 (2) | 20 (2) | 26 (2) |  |  |
| Undetermined | 10 (0.8) | 15 (1) | 20 (2) |  |  |
| Therapeutic complication | 2 (0.2) | 1 (0.08) | 0 |  |  |
| **Substances present<sup>b</sup>** |  |  |  |  |  |
| Heroin | 341 (26) | 247 (19) | 157 (12) | <0.001*** | <0.001*** |
| Alcohol | 250 (19) | 299 (23) | 350 (26) | 0.02* | 0.04* |
| Amphetamines<sup>d</sup> | 43 (3) | 61 (5) | 113 (9) | 0.08 | <0.001*** |
| Fentanyl analogs | 82 (6) | 117 (9) | 53 (4) | 0.01* | <0.001*** |
| Fentanyl | 1113 (85) | 1162 (90) | 1171 (89) | 0.001** | 0.51 |
| Benzodiazepines | 362 (28) | 292 (22) | 297 (22) | 0.002** | >0.99 |
| Prescription opioids<sup>c</sup> | 128 (10) | 112 (9) | 137 (10) | 0.33 | 0.15 |
| Cocaine | 485 (37) | 528 (41) | 585 (44) | 0.07 | 0.07 |
| **Substances with and without fentanyl** |  |  |  |  |  |
| Benzodiazepines with fentanyl | 304 (23) | 259 (20) | 255 (19) | 0.04* | 0.71 |
| Benzodiazepines without fentanyl | 58 (4) | 33 (3) | 42 (3) | 0.01* | 0.39 |
| Cocaine or amphetamines with fentanyl | 412 (32) | 493 (38) | 554 (42) | <0.001*** | 0.04* |
| Cocaine or amphetamines without fentanyl | 98 (8) | 76 (6) | 99 (7) | 0.11 | 0.11 |
| Alcohol with fentanyl | 210 (16) | 263 (20) | 316 (24) | 0.007** | 0.03* |
| Alcohol without fentanyl | 40 (3) | 36 (3) | 34 (3) | 0.74 | 0.84 |

<sup>a</sup>Significance: \*<0.05; \*\*<0.01; \*\*\*<0.001; <sup>b</sup>Categories are not mutually exclusive because deaths can involve multiple drugs;
<sup>c</sup>Includes hydrocodone, hydromorphone, oxycodone, oxymorphone, codeine, dihydrocodeine, levorphanol and tramadol;
<sup>d</sup>Includes methamphetamine.

On average, there were 5.7 (95% CI: 5.4 to 6.0) OD deaths per day in 2018, 5.7 (95% CI: 5.4 to 6.0) in 2019 and 5.8 (95% CI: 5.5 to 6.1) in 2020; there was no evidence that the change in 2020 was significant (P=0.61).

Across all three periods, there was no evidence of a significant change in the sex distribution of OD decedents. From 2019 to 2020, there was a 2.6% increase in the average age of OD decedents (P=0.02). In 2019, 78% of OD decedents were White, vs. 73% in 2020; this decrease contrasts an increase in Black decedents from 7% to 10% (P=0.02). From 2018 to 2019, there was no evidence of a significant change in the distribution of decedents’ race (P=0.10). There was no evidence of significant changes in the distribution of manner of death from 2018 to 2019 (P=0.70) or from 2019 to 2020 (P=0.50).

From 2018 to 2020, heroin’s presence consistently decreased (26% in 2018, 19% in 2019, and 12% in 2020, P<0.001]), and alcohol’s presence increased (19% in 2018 vs. 23% in 2019 [P=0.02] and 26% in 2020 [P=0.04]). Changes in trends were observed for amphetamines, with no evidence for changes in its presence from 2018 to 2019 (3% vs. 5%, P=0.08) but increased presence from 2019 to 2020 (5% vs. 9%, P<0.001). Changes were also observed for fentanyl analogs, which increased from 2018 to 2019 (6% vs. 9%, P=0.01) but decreased from 2019 to 2020 (9% vs. 4%, P<0.001). From 2018 to 2019, fentanyl presence increased (85% vs. 90%, P=0.001), but it remained steady from 2019 to 2020 (90% vs. 89%; P=0.51). Benzodiazepine presence decreased from 2018 to 2019 (28% vs. 22%, P=0.002), but was constant from 2019 to 2020 (22%, P>0.99). Prescription opioid presence was constant from 2019 to 2020 (9% vs. 10%; P=0.15), as was cocaine presence (41% vs. 44%, P=0.15).

Benzodiazepine-involved OD deaths both with and without fentanyl decreased from 2018 to 2019 (with fentanyl: 23% vs. 20%, P=0.04; without fentanyl: 4% vs. 3%, P=0.01) and were steady from 2019 to 2020 (with fentanyl: 20% vs. 19%, P=0.71; without fentanyl: 3% vs. 3%, P=0.39). OD deaths involving cocaine or amphetamines and fentanyl increased from 2018 to 2019 (32% vs. 38%, P<0.001) and from 2019 to 2020 (38% vs. 42%, P=0.04), as did OD deaths involving alcohol and fentanyl (16% in 2018, 20% in 2019 [P=0.007], and 24% in 2020 [P=0.03]). OD deaths involving cocaine or amphetamines without fentanyl remained steady across both periods (P=0.11), as did deaths involving alcohol without fentanyl, which constituted 3% of deaths in all three years.

We found no evidence of significant correlation between the COVID-19 case fatality rates and the percentage change in OD deaths at the county level from 2019 to 2020 (r=0.34, 95% CI: - 0.29 to 0.77).

## Discussion

During COVID-19, the drug overdose crisis has remained a formidable foe to public health. From 2019 to 2020, there were critical signals of increased amphetamine-involved ODs and ODs involving cocaine or amphetamines with fentanyl and alcohol with fentanyl in Massachusetts, which aligns with national-level increases in concurrent stimulant- and opioid-involved overdoses and overdose deaths (Hoots, Vivolo-Kantor, & Seth, 2020). Additionally, the signals of increased alcohol presence in drug ODs corroborate reports of increased alcohol consumption over the course of the pandemic (Pollard et al., 2020), a trend which was also present prior to the pandemic (Warren, 2021). Following prior trends, the proportion of ODs involving heroin decreased. Notably, fentanyl was present in more than 85% of ODs during all three periods, reflecting its continued domination of the illicit opioid supply, especially in Massachusetts, and increasing rates of fentanyl use with or without other substances.

Further, the increase in the proportion of Black OD decedents mirrors recent reports of increasing overdose deaths among Black people (James & Jordan, 2018), which had begun to materialize before the onset of COVID-19. Systemic racism evident in decreased access to health care, safe occupational conditions, income and wealth, and educational opportunities, present prior to the overdose crisis’ rise and certainly prior to the COVID-19 pandemic, may have been exacerbated by the compounding crises; however, extensions to this research are needed to characterize the causal effects at the intersection of COVID-19 and the overdose crisis.

Recent research (Glober et al., 2020; Rodda et al., 2020) indicates increased daily opioid-related mortality following the pandemic’s onset. Additionally, there are reports of increased initiation and use of substances to cope with the impacts of COVID-19 (Czeisler et al., 2020). We observed a steady trend in daily drug ODs in Massachusetts, which is nonetheless notable given that the state was finally beginning to see a decline in overdose fatality rates prior to COVID-19 (National Institute on Drug Abuse, 2020).

In light of inconclusive evidence for an increase in OD trends across the state, we investigated whether there was a relationship between COVID-19 severity and OD deaths at the county level. This analysis revealed no evidence that COVID-19 case fatality rate and increase in OD death rates are correlated.

Our analysis does not provide insight into the reason behind Massachusetts’ steady OD rate, in contrast to other states’ OD surges and the overall national increase, but several policy choices in Massachusetts merit further investigation. First, Massachusetts took several steps to ensure ongoing access to treatment and harm reduction resources, which may explain the non-significant increase in fatal ODs. For example, the state increased access to medications for opioid use disorder and issued guidance for OD rescue during the pandemic, potentially reducing OD deaths (Massachusetts Department of Public Health, 2020). In contrast, California, which observed increases in OD deaths, did not enact statewide methadone expansion in concordance with SAMHSA’s March 2020 guidelines. Further, California already had increasing numbers of OD deaths, beginning in mid-2019, as opposed to the relatively steady trend in ODs observed in Massachusetts from mid-2018 to May 2020 (National Center for Health Statistics, 2021). Indiana has also experienced varying OD death trends over the last three years; deaths decreased from 2018 to 2019, and increased from 2019 to May 2020 (National Center for Health Statistics, 2021). Indiana did implement methadone access expansion in March 2020; however, Indiana was among at least five states (along with Florida, Illinois, Pennsylvania, and Minnesota) in which opioid treatment facilities were forced to shut down as a result of the pandemic (Bruce, 2020). There are myriad factors that may contribute to heterogenous OD trends among states. The source of differences between findings in various states is critical and warrants careful investigation in future research.

This analysis has limitations, chiefly that it is a purely correlative analysis which cannot provide insight into the causes of the identified trends. Whether COVID-19 caused the demographic or substance shifts in overdose deaths in Massachusetts is subject to further investigation; this analysis may form the basis of future causal analyses. In addition, our findings only reflect trends through November 8, 2020. Policies to mitigate the effects of COVID-19 are informed by detailed real-time infection and mortality data. Drug overdose data, however, are collated more slowly than COVID-19 data; state data are finalized on a three- to four-month lag in Massachusetts and these lags are often greater in other states. Our ongoing work aims to link several up-to-date data sources to better characterize and forecast the impacts of COVID-19 on the overdose crisis.

Analysis of available data reveals signals of increased amphetamines and alcohol presence in ODs in Massachusetts. Amphetamine-related and combined stimulant and fentanyl-related deaths were becoming more frequent prior to the pandemic, but COVID-19 may have exacerbated their rise. Alcohol use has also increased, and the interaction between this and existing opioid and stimulant use may amplify already-dangerous trends in ODs. The risk of stimulant, alcohol, and benzodiazepine-related overdoses are exacerbated with increasing use and co-use of fentanyl in addition to its continued contamination of illicit opioids in Massachusetts, among other states. This underscores the need for rapid and low-threshold access to effective medication treatment, psychosocial support, and harm reduction resources. Prevention, treatment, and harm reduction interventions must be rooted in a race equity framework and address the broader contextual factors of homelessness, lost economic opportunity, and societal despair while accounting for the increasing role of fentanyl in combination with stimulants and alcohol in the overdose crisis.

## Data Availability

Data will be available upon request.

## Author Contributions

C.D. and G.G.P.G. collected the data and performed statistical analysis and interpretation of data. E.S., S.W., and M.S.J. provided critical review of the analysis and contributed to further development of the content. C.D. and G.G.P.G. drafted the manuscript and E.S., S.W., and M.S.J. revised the manuscript for important intellectual content. M.S.J. conceived the study and supervised the project. All authors read and approved the final manuscript.

## Acknowledgments

We thank Rosalie Pacula for providing critical feedback on the analysis. We also thank Elizabeth Beaulieu, Huiru Dong, Simin Falsafi, Nicole Poellinger, and Celia Stafford for sharing their constructive comments on earlier versions of this manuscript. No funding was used to conduct this analysis.

